# Acceptability of human papillomavirus (HPV) self-sampling among never- and under-screened Indigenous and other minority women: a randomised three-arm community trial in Aotearoa New Zealand

**DOI:** 10.1101/2021.04.11.21255231

**Authors:** Naomi Brewer, Karen Bartholomew, Jane Grant, Anna Maxwell, Georgina McPherson, Helen Wihongi, Collette Bromhead, Nina Scott, Sue Crengle, Sunia Foliaki, Chris Cunningham, Jeroen Douwes, John D. Potter

## Abstract

**Background:** Internationally, self-sampling for human papillomavirus (HPV) has been shown to increase participation in cervical-cancer screening. In Aotearoa New Zealand, there are long-standing ethnic inequalities in cervical-cancer screening, incidence, and mortality; particularly for indigenous Māori women, as well as Pacific, and Asian women.

**Methods:** We invited never- and markedly under-screened (≥5 years overdue) 30-69-year-old Māori, Pacific, and Asian women to participate in an open-label, three-arm, community-based, randomised controlled trial, with a nested sub-study. We aimed to assess whether two specific invitation methods for self-sampling improved screening participation over usual care among the least medically served populations. Women were individually randomised 3:3:1 to: clinic-based self-sampling (CLINIC – invited to take a self-sample at their usual general practice); home-based self-sampling (HOME – mailed a kit and invited to take a self-sample at home); and usual care (USUAL – invited to attend a clinic for collection of a standard cytology sample). Neither participants nor research staff could be blinded to the intervention. In a subset of general practices, women who did not participate within three months of invitation were opportunistically invited to take a self-sample, either next time they attended a clinic or by mail.

**Findings:** We randomised 3,553 women: 1,574 to CLINIC, 1,467 to HOME, and 512 to USUAL. Participation was highest in HOME (14.6% among Māori, 8.8% among Pacific, and 18.5% among Asian) with CLINIC (7.0%, 5.3% and 6.9%, respectively) and USUAL (2.0%, 1.7% and 4.5%, respectively) being lower. In fully adjusted models, participation was statistically significantly more likely in HOME than USUAL: Māori OR=9.7, (95%CI 3.0-31.5); Pacific OR=6.0 (1.8-19.5); and Asian OR=5.1 (2.4-10.9). There were no adverse outcomes reported. After three months, 2,780 non-responding women were invited to participate in a non-randomised, opportunistic, follow-on substudy. After 6 months,192 (6.9%) additional women had taken a self-sample.

**Interpretation:** Using recruitment methods that mimic usual practice, we provide critical evidence that self-sampling increases screening among the groups of women (never and under-screened) who experience the most barriers in Aotearoa New Zealand, although the absolute level of participation through this population approach was modest. Follow-up for most women was routine but a small proportion required intensive support.

**Trial registration:** ANZCTR Identifier: ACTRN12618000367246 (date registered 12/3/2018) https://www.anzctr.org.au/Trial/Registration/TrialReview.aspx?id=371741&isReview=true;UTN:U1111-1189-0531

**Funding:** Health Research Council of New Zealand (HRC 16/405)

**Protocol:** http://publichealth.massey.ac.nz/assets/Uploads/Study-protocol-V2.1Self-sampling-for-HPV-screening-a-community-trial.pdf

## Background

Internationally, the accuracy of human papillomavirus (HPV) self-sampling for cervical-cancer screening has been shown to be similar to healthcare-professional-taken samples when tested for high-risk (hr) HPVs using polymerase chain reaction (PCR)^1-5^. Offering self-sampling improves participation^6^, with differences in that improvement shown when it is offered at a population level (*i*.*e*., mail-out to all potentially eligible women) compared to when it is offered at the individual level (*i*.*e*., face-to-face)^2^. However, most studies have been targeted at the dominant population group or socioeconomically deprived populations and not Indigenous or ethnic-minority groups, who are underserved in many health systems^7,8^.

In Aotearoa New Zealand, the success of the National Cervical Screening Programme (NCSP) has not been equitable for Māori (the Indigenous people of Aotearoa New Zealand), Pacific, and Asian women. Māori and Pacific women have higher cervical cancer incidence and mortality rates than European/Other women^9^ and lower screening rates^9^. Asian women also have lower screening rates^9^, although incidence is lower among Asian women than European/Other women and mortality rates are similar^9^. Although the women who do not participate in screening are often referred to as ‘hard-to-reach’ or ‘disengaged’, we consider the inequities a systems issue and therefore refer to these groups as underserved.

The majority of cervical cancer cases in Aotearoa New Zealand occur in women who have never been screened or are under-screened^10,11^. There have been a variety of measures introduced to increase screening rates, but these have underperformed. The longstanding nature of the problem shows that novel strategies are needed. As Aotearoa New Zealand transitions to primary HPV-based screening, there is an opportunity to introduce self-sampling.

The World Health Organization’s strategy to accelerate the elimination of cervical cancer notes the need to examine screening implementation in the local context^12^. One recent study of self-testing in Aotearoa New Zealand assessed the impact of intense recruitment of all overdue women^13^. In contrast, our approach concentrated on the least served high-priority populations of never- and under-screened Māori, Pacific, and Asian women and was designed to reflect how self-sampling will probably be implemented in Aotearoa New Zealand. We aimed to evaluate whether two specific invitation methods for self-sampling could increase screening participation. We also sought to obtain information on the resources required to achieve 90% follow-up of hrHPV-positive women. Our study is the first to evaluate the effectiveness of a mailed self-sampling kit for cervical-cancer screening in Aotearoa New Zealand.

## Methods

We have published the study objectives and methods (including the exclusion criteria and the main study interventions) previously^14^. Briefly, this was an open-label, three-arm, community-based, randomised controlled trial, with a non-randomised follow-on sub-study. We invited never- and under-screened (no screening recorded for at least the last five years^15^) 30-69 year-old Māori, Pacific, and Asian women in the Auckland area (Waitematā and Auckland District Health Boards (DHBs)).

Participating clinics were selected because they had enrolled populations with high proportions of Māori, Pacific, and Asian women who were never- or under-screened. Clinics were also selected to represent a range of small and large clinics.

Once participants reached the status of being eligible for the study, the study programmer had the system i) count the number of patients already assigned to each arm and ii) randomly allocate the next patient to maintain the required ratio across the three arms.

Women were individually randomised 3:3:1 to: clinic-based self-sampling, in which women were invited to take a self-sample at their usual general practice (GP); home-based self-sampling, in which women were mailed a kit and invited to take a self-sample at home; and usual care, in which women were offered standard cytology (at a clinic, at an independent service provider, or with the study nurse). Women were directed to the study webpage with translated study documents and study video clips (with subtitles in Te Reo Māori, Tongan, Samoan, Korean, and Simplified Chinese). Participation was confirmed through the NCSP-Register, GP records, or receipt of a sample at the lab. All tests were without charge. Prior to joining the study, clinic staff were trained in study procedures and returning results to women.

Women in the self-sampling arms were given a FLOQSwab™ (Copan Italia, Brescia, Italy), with which to take a low-vaginal sample and a 12 mL dry tube (Sarstedt AG & Co. KG, Germany) for sample transportation. HPV testing was carried out with the cobas 4800 HPV assay (Roche Molecular Systems, Pleasanton, California, USA) at a single laboratory: Anatomic Pathology Service (the International Accreditation New Zealand (IANZ) Accredited laboratory of Auckland DHB).

### Non-randomised follow-on sub-study invitation

In a subset of clinics, women (including those originally randomised to usual care) who had not responded three months after their invitation were sent a letter or text message informing them that they could self-sample at their clinic or request that a self-sampling kit be mailed to them, both for a limited period of time (6 or 9 months; the difference was due to study-duration limits). Women were able to take the sample at the clinic or take the kit home.

### Sample size and power

We aimed to invite a minimum of 3,550 un- or under-screened Māori, Pacific, and Asian women (1,050, 1,250, 1,250 respectively). With 450 Māori women invited in the clinic self-sampling group and 150 Māori women invited to usual-care, we would have >85% power to detect a 10% difference in uptake between groups (*e*.*g*., 15% uptake in the clinic group and 5% usual-care)^14^.

### Results management

Negative results were provided to women by the usual primary-care provider and, with agreement from the NCSP, women were advised to return for a routine cervical screen at the appropriate clinical interval (3-year recall interval)^15^, as specified by the NCSP guidelines and an amended approach to the proposed HPV primary-screening algorithm.

Women with inadequate or abnormal cytology were followed-up according to the NCSP guidelines^15^ by the requesting smear-taker.

Positive HPV results were managed as per the NCSP guidelines, with adjustments in accordance with NCSP clinical advice^16^. Women who tested HPV16/18 positive were referred directly to colposcopy. Women who tested positive for any of the pool of 12 other hrHPV types were triaged with a clinician-conducted cervical-cytology test; however, if a woman declined cytology, she was offered colposcopy to ensure safe follow-up.

### Statistical analysis

The main analyses focused on the primary outcome of the study: participation, *i*.*e*. the proportion of women who provided a self-sample compared with the proportion who attended for cytology, stratified by ethnicity. These data were initially assessed simply by comparing (with Pearson’s *x*^2^-tests) the proportions who participated in each group and by logistic regression models to adjust for potential confounders (study group, age group, screening history, and socioeconomic status (assessed through a measure of small area relative deprivation; the New Zealand Deprivation Index 2018 (NZDep)^17^). We report here the intention-to-treat analyses.

The laboratory turnaround time (date received to date reported) and proportion of unsatisfactory samples was monitored.

Statistical analyses were done using Stata 15.1 (Stata Corp LLC, College Station, TX, USA).

### Ethics requirements

The study was approved by the New Zealand Northern B Health and Disability Ethics Committee (HDEC) (reference: 17/NTB/120). The New Zealand Ministry of Health (including the National Kaitiaki Group (a group of Māori women established to monitor the safe use of Māori data within the NCSP)) and the participating DHBs, primary health organisations and clinics approved the use of data to identify and contact eligible women.

## Results

Twenty-three clinics agreed to participate, through which we identified 5,546 women who were initially considered eligible for the study. Of these, 853 (853/5,546, 15%) were excluded prior to being sent a pre-invitation (Figure 1 and Supplementary Table 1). Recruitment began 8^th^ June 2018. After being sent the pre-invitation, a further 128 (128/4,693, 3%) women were excluded and 37 opted out. A further 900 women (900/4,528, 20%) were excluded after being randomised and 75 opted out, leaving 3,553 randomised women. Exclusions were proportionally similar by ethnicity for the major contributors of hysterectomy, pregnancy, and symptoms; however exclusions for previous high-grade cytology were higher for Māori (60%) than for Pacific (26%) and Asian women (14%). Recruitment ended 13^th^ May 2020.

**Figure 1.**
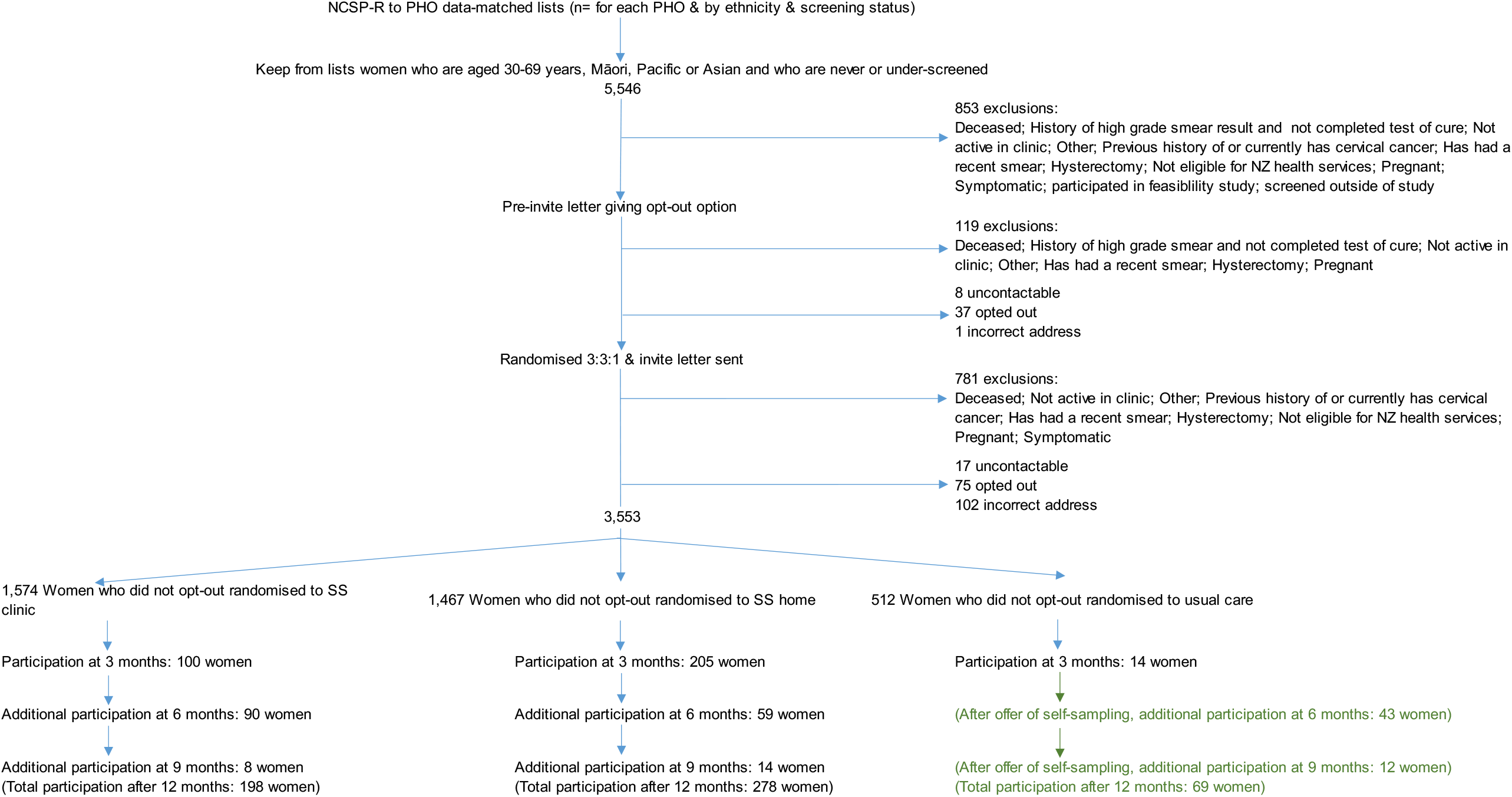
CONSORT flowchart of participation. NCSP-R: National Cervical Screening Programme-Register; NZ: Aotearoa New Zealand; PHO: Primary Health Organisation; SS: self-sampling.

The median age of the participants was 44 years (interquartile range 36-55 years; Table 1). Slightly fewer Māori women (1,073) were randomised than Pacific (1,251) and Asian (1,229) women and a slightly higher proportion of Asian women (68%) were in the youngest two age categories (30-39 and 40-49 years) than Māori (63%) and Pacific (64%) women. A higher proportion of never-screened women (72%) than under-screened women (60%) were in the youngest two age categories and more Asian women (68%) than Māori (22%) and Pacific (39%) women were never screened. Overall, approximately 60% of the women lived in areas of high socioeconomic deprivation (quintiles four and five).

**Table 1:** Participant demographics.

|  | Overall<br>n (%) | Ethnic group |  |  | Previous screening history |  |
| --- | --- | --- | --- | --- | --- | --- |
|  |  | Māori<br>n (%) | Pacific<br>n (%) | Asian<br>n (%) | Never screened<br>n (%) | Under-screened<br>n (%) |
| **Median age** | 44.0 | 45.1 | 45.0 | 41.9 | 40.0 | 46.5 |
| **Age group** |  |  |  |  |  |  |
| 30-39 | 1,352 (38.1) | 364 (33.9) | 438 (35.0) | 550 (44.8) | 783 (50.1) | 569 (28.6) |
| 40-49 | 955 (26.9) | 310 (28.9) | 356 (28.5) | 289 (23.5) | 339 (21.7) | 616 (31.0) |
| 50-59 | 759 (21.4) | 258 (24.0) | 306 (24.5) | 195 (15.9) | 231 (14.9) | 528 (26.5) |
| 60-69 | 487 (13.7) | 141 (13.1) | 151 (12.1) | 195 (15.9) | 210 (13.4) | 277 (13.9) |
| **Ethnicity** |  |  |  |  |  |  |
| Māori | 1,073 (30.2) | - | - | - | 234 (21.8) | 839 (78.2) |
| Pacific | 1,251 (35.2) | - | - | - | 490 (39.2) | 761 (60.8) |
| Asian | 1,229 (34.6) | - | - | - | 839 (68.3) | 390 (31.7) |
| **Previous screening history** |  |  |  |  |  |  |
| Never screened | 1,563 (44.0) | 234 (21.8) | 490 (39.2) | 839 (68.3) | - | - |
| Under-screened | 1,990 (56.0) | 839 (78.2) | 761 (60.8) | 390 (31.7) | - | - |
| **NZDep2018, quintile** |  |  |  |  |  |  |
| 1 (least deprived) | 270 (7.6) | 62 (5.8) | 44 (3.5) | 164 (13.3) | 131 (8.4) | 139 (7.0) |
| 2 | 466 (13.1) | 126 (11.7) | 90 (7.2) | 250 (20.3) | 227 (14.5) | 239 (12.0) |
| 3 | 679 (19.1) | 173 (16.1) | 205 (16.4) | 301 (24.5) | 322 (20.7) | 357 (17.9) |
| 4 | 959 (27.0) | 295 (27.5) | 351 (28.1) | 313 (25.5) | 424 (27.1) | 535 (26.9) |
| 5 (most deprived) | 1,178 (33.2) | 417 (38.9) | 560 (44.8) | 201 (16.4) | 458 (29.3) | 720 (36.2) |
| Missing | 1 (0.0) | 0 | 1 (0.1) | 0 | 1 (0.1) | 0 |

Column percentages add to 100% (apart from rounding). NZDep: New Zealand Deprivation Index 2018.

The highest participation proportions for the initial three-month period (the randomised phase) were in the home-based group (14.0%, p <0.0001 compared to usual care); compared with 6.4% in the clinic-based group and 2.7% in the usual-care group (Table 2). Participation was higher in the home-based group than either the clinic-based or usual-care groups across all of the demographic variables (although the differences were not always statistically significant). The pattern was similar for the clinic-based group in comparison to the usual-care group. Māori (14.6%) and Asian (18.5%) women had greater participation than Pacific women (8.8%) in the home-based group. Participation proportions generally decreased as deprivation level increased in the clinic-based and home-based groups, but this pattern was not seen in the usual-care group.

**Table 2:** Participation proportions during main randomised phase (90 days from invite)

|  | Randomised group |  |  | Took a sample |  |  | Participation proportion |  |  |  |  |  |  |
| --- | --- | --- | --- | --- | --- | --- | --- | --- | --- | --- | --- | --- | --- |
|  | Clinic-based self-sampling n (%) | Home-based self-sampling n (%) | Usual care n (%) | Clinic-based self-sampling n | Home-based self-sampling n | Usual care n | Clinic-based self-sampling % | Clinic Pearson chi2 'versus' usual care p | Home-based self-sampling % | Home Pearson chi2 'versus' usual care p | Self-sampling groups combined % | Combined Pearson chi2 'versus' usual care p | Usual care % |
| **All** | 1574 (44.3) | 1467 (41.3) | 512 (14.4) | 100 | 205 | 14 | 6.4 | 0.002 | 14.0 | 0.000 | 10.0 | 0.000 | 2.7 |
| **Age group** |  |  |  |  |  |  |  |  |  |  |  |  |  |
| 30-39 | 578 (36.7) | 577 (39.3) | 197 (38.5) | 29 | 87 | 5 | 5.0 | 0.142 | 15.1 | 0.000 | 10.0 | 0.001 | 2.5 |
| 40-49 | 434 (27.6) | 376 (25.6) | 145 (28.3) | 35 | 48 | 5 | 8.1 | 0.058 | 12.8 | 0.002 | 10.2 | 0.009 | 3.4 |
| 50-59 | 341 (21.7) | 311 (21.2) | 107 (20.9) | 21 | 40 | 1 | 6.2 | 0.029 | 12.9 | 0.000 | 9.4 | 0.003 | 0.9 |
| 60-69 | 221 (14.0) | 203 (13.8) | 63 (12.3) | 15 | 30 | 3 | 6.8 | 0.561 | 14.8 | 0.035 | 10.6 | 0.146 | 4.8 |
| **Ethnicity** |  |  |  |  |  |  |  |  |  |  |  |  |  |
| Māori | 469 (29.8) | 451 (30.7) | 153 (29.9) | 33 | 66 | 3 | 7.0 | 0.020 | 14.6 | 0.000 | 10.8 | 0.001 | 2.0 |
| Pacific | 569 (36.1) | 502 (34.2) | 180 (35.2) | 30 | 44 | 3 | 5.3 | 0.040 | 8.8 | 0.001 | 6.9 | 0.007 | 1.7 |
| Asian | 536 (34.1) | 514 (35.0) | 179 (35.0) | 37 | 95 | 8 | 6.9 | 0.246 | 18.5 | 0.000 | 12.6 | 0.002 | 4.5 |
| **Screening history** |  |  |  |  |  |  |  |  |  |  |  |  |  |
| Never screened | 704 (44.7) | 657 (44.8) | 202 (36.5) | 44 | 96 | 6 | 6.3 | 0.072 | 14.6 | 0.000 | 10.3 | 0.001 | 3.0 |
| Under screened | 870 (55.3) | 810 (55.2) | 310 (60.5) | 56 | 109 | 8 | 6.4 | 0.010 | 13.5 | 0.000 | 9.8 | 0.000 | 2.6 |
| **Ethnicity & Screening history** |  |  |  |  |  |  |  |  |  |  |  |  |  |
| Māori never screened | 109 (23.2) | 101 (22.4) | 24 (15.7) | 8 | 13 | 0 | 7.3 | 0.171 | 12.9 | 0.063 | 10.0 | 0.104 | 0.0 |
| Māori under-screened | 360 (76.8) | 350 (77.6) | 129 (84.3) | 25 | 53 | 3 | 6.9 | 0.053 | 15.1 | 0.000 | 11.0 | 0.002 | 2.3 |
| Pacific never screened | 236 (41.5) | 192 (38.3) | 62 (34.4) | 12 | 17 | 2 | 5.1 | 0.538 | 8.9 | 0.143 | 6.8 | 0.283 | 3.2 |
| Pacific under-screened | 333 (58.5) | 310 (61.8) | 118 (65.6) | 18 | 27 | 1 | 5.4 | 0.034 | 8.7 | 0.003 | 7.0 | 0.010 | 0.8 |
| Asian never screened | 359 (66.9) | 364 (70.8) | 116 (64.8) | 24 | 66 | 4 | 6.7 | 0.198 | 18.1 | 0.000 | 12.4 | 0.004 | 3.4 |
| Asian under-screened | 177 (33.0) | 150 (29.2) | 63 (35.2) | 13 | 29 | 4 | 7.3 | 0.791 | 19.3 | 0.017 | 12.8 | 0.143 | 6.3 |
| **NZDep2018, quintile** |  |  |  |  |  |  |  |  |  |  |  |  |  |
| 1 (least deprived) | 132 (8.4) | 105 (7.2) | 33 (6.4) | 8 | 23 | 2 | 6.1 | 1.000 | 21.9 | 0.039 | 13.1 | 0.249 | 6.1 |
| 2 | 203 (12.9) | 188 (12.8) | 75 (14.6) | 18 | 38 | 1 | 8.9 | 0.027 | 20.2 | 0.000 | 14.3 | 0.002 | 1.3 |
| 3 | 287 (18.2) | 290 (19.8) | 102 (19.9) | 22 | 44 | 2 | 7.7 | 0.040 | 15.2 | 0.000 | 11.4 | 0.003 | 2.0 |
| 4 | 437 (27.8) | 387 (26.4) | 135 (26.4) | 32 | 57 | 4 | 7.3 | 0.068 | 14.7 | 0.000 | 10.8 | 0.004 | 3.0 |
| 5 (most deprived) | 515 (32.7) | 497 (33.9) | 166 (32.4) | 20 | 43 | 5 | 3.9 | 0.604 | 8.7 | 0.015 | 6.2 | 0.100 | 3.0 |
| Missing | 0 | 0 | 1 (0.2) | 0 | 0 | 0 | - | - | - | - | - | - | - |

Column percentages add to 100% (apart from rounding). P value: Pearson's $\chi^2$ vs. usual care. NZDep: New Zealand Deprivation Index 2018.

Figure 2 shows the proportion of women screened up to 90 days after invitation (randomised phase). Women randomised to the home-based group appeared to take a sample earlier after receiving an invitation than women who were randomised to clinic-based and usual-care groups (Figure 2a).

**Figure 2:**
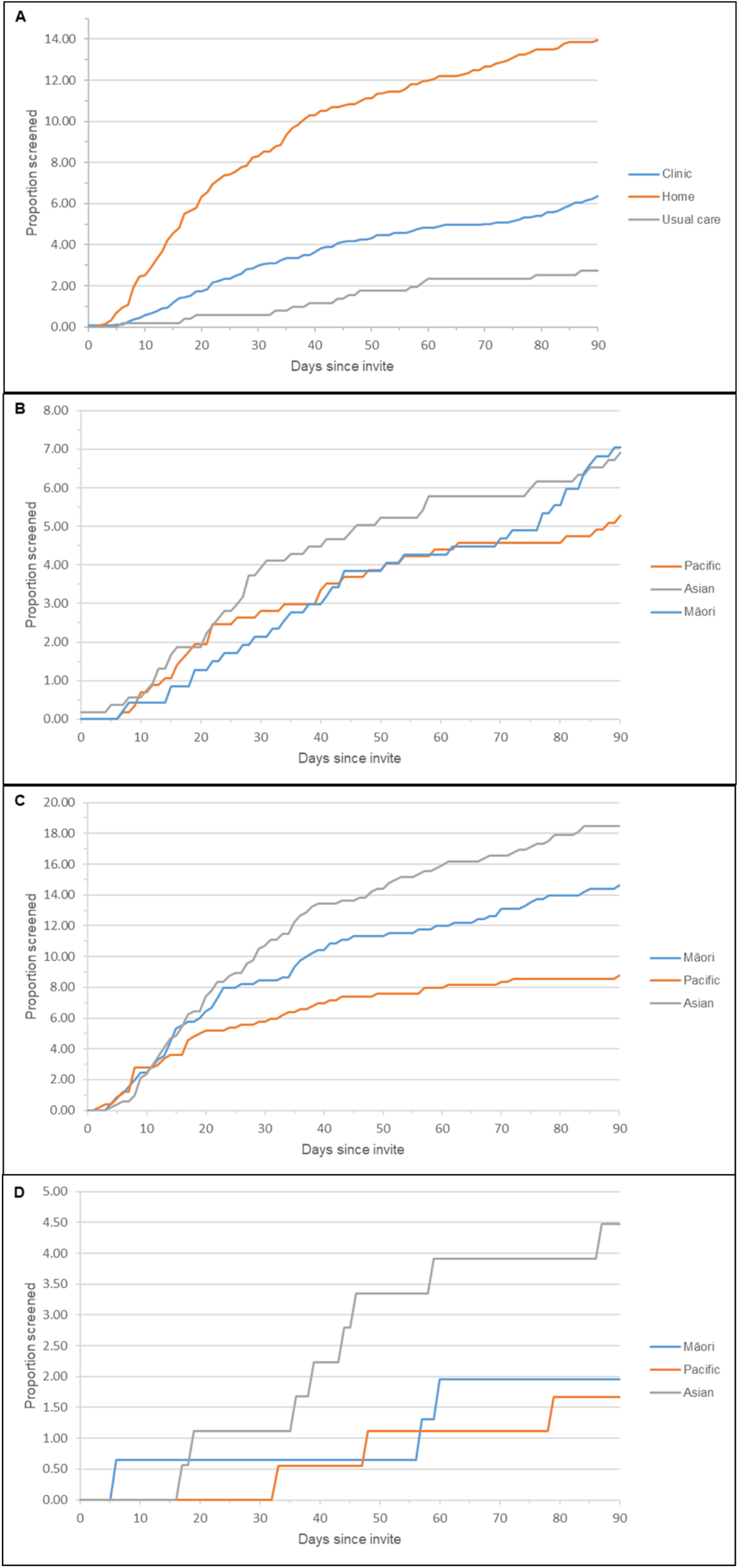
Participation proportions during main randomised phase (90 days from invite) A: Participation proportion by randomised group; B: Participation in clinic group; C: Participation in home-based group; D: Participation in usual care group

Previous screening history did not have a large impact on participation: among the never-screened, home-based participation was 14.6% and under-screened participation was 13.5%; clinic participation was 6.3% and 6.4%, respectively; and usual-care participation 3.0% and 2.6% respectively (Table 2).

Unadjusted regression stratified by ethnicity showed that participation in the three-month randomised phase was statistically significantly higher in the home-based group than in the usual-care group (Māori: OR 8.6; 95%CI 2.7-27.7; Pacific: 5.7; 1.7-18.5; and Asian: 4.9; 2.3-10.2; Table 3), although the confidence intervals were wide. The ORs remained largely unchanged – and statistically significant – in the fully adjusted model. Māori women aged 60-69 years had more than twice the odds of participation (2.6; 1.4-4.6) of women aged 30-39 years, whereas Asian women aged 60-69 years had lower odds (0.5; 0.3-1.0). Asian women in NZDep quintile 4 had lower odds of participation (0.5; 0.3-0.9) than Asian women living in the least-deprived area. Again, these estimates remained largely unchanged in the fully adjusted models (Table 3).

**Table 3:** Odds of participation during the randomised phase (up to 90 days after invitation)

|  | Odds ratio (95% CI) of participation |  |  |  |  |  |
| --- | --- | --- | --- | --- | --- | --- |
|  | Unadjusted |  |  | Adjusted for study group, screening history, NZDep & age group |  |  |
|  | Māori | Pacific | Asian | Māori | Pacific | Asian |
| Study group |  |  |  |  |  |  |
| Clinic | 3.8 (1.1-12.5) | 3.3 (1.0-10.9) | 1.6 (0.7-3.5) | 4.1 (1.2-13.5) | 3.3 (1.0-11.0) | 1.6 (0.7-3.5) |
| Home | 8.6 (2.7-27.7) | 5.7 (1.7-18.5) | 4.9 (2.3-10.2) | 9.7 (3.0-31.5) | 6.0 (1.8-19.5) | 5.1 (2.4-10.9) |
| Usual care | Ref | Ref | Ref | Ref | Ref | Ref |
| Age group |  |  |  |  |  |  |
| Age 30-39 | Ref | Ref | Ref | Ref | Ref | Ref |
| Age 40-49 | 1.4 (0.8-2.4) | 1.25 (0.7-2.2) | 0.9 (0.6-1.4) | 1.3 (0.8-2.3) | 1.3 (0.7-2.4) | 0.9 (0.5-1.4) |
| Age 50-59 | 1.1 (0.6-1.9) | 1.1 (0.6-2.1) | 0.9 (0.6-1.5) | 1.0 (0.5-1.8) | 1.2 (0.7-2.4) | 0.8 (0.5-1.3) |
| Age 60-69 | 2.6 (1.4-4.6) | 1.2 (0.6-2.6) | 0.5 (0.3-1.0) | 2.6 (1.4-4.7) | 1.2 (0.6-2.6) | 0.5 (0.3-0.9) |
| Screening history |  |  |  |  |  |  |
| Never screened | 0.9 (0.6-1.5) | 1.1 (0.7-1.7) | 0.9 (0.7-1.4) | 0.9 (0.5-1.6) | 1.1 (0.7-1.7) | 0.9 (0.6-1.3) |
| Under screened | Ref | Ref | Ref | Ref | Ref | Ref |
| NZDep2018, quintile |  |  |  |  |  |  |
| 1 (least deprived) | Ref | Ref | Ref | Ref | Ref | Ref |
| 2 | 1.4 (0.5-4.2) | 1.0 (0.2-4.1) | 1.0 (0.6-1.7) | 1.8 (0.6-5.5) | 0.9 (0.2-3.9) | 0.9 (0.5-1.7) |
| 3 | 1.8 (0.7-5.1) | 0.7 (0.2-2.7) | 0.7 (0.4-1.2) | 2.2 (0.8-6.2) | 0.7 (0.2-2.5) | 0.6 (0.3-1.1) |
| 4 | 1.7 (0.6-4.5) | 1.3 (0.4-4.4) | 0.5 (0.3-0.9) | 2.1 (0.8-5.5) | 1.3 (0.4-4.3) | 0.4 (0.2-0.8) |
| 5 (most deprived) | 0.6 (0.2-1.7) | 0.7 (0.2-2.5) | 0.6 (0.3-1.1) | 0.7 (0.3-2.0) | 0.7 (0.2-2.3) | 0.5 (0.3-1.0) |

95%CI: 95% confidence interval; NZDep: New Zealand Deprivation Index 2018.

There were no adverse outcomes reported.

In order to test whether the setting of a formal clinical trial provides only a floor for the estimate of acceptability in this underserved population, 2,780 non-responding women (1,284 clinic, 1,070 home-based, and 426 usual-care) were invited to participate in a non-randomised follow-on substudy involving only the opportunity to self-sample. After 6 months,192 (6.9%: 7.0% clinic-group; 5.5% home-based; and 10.1% usual-care) had taken a self-sample. The clinics that started recruitment early in the study facilitated opportunistic participation up to 9 months and this yielded a further 34 self-samples, again with a somewhat higher uptake among those originally randomised to usual-care (7.5%) than among the two self-sampling arms (2.4%). Participation in this substudy was censored on 25^th^ March 2020 when Aotearoa New Zealand went into a strict COVID-19-related lockdown; we reasoned that women would not easily be able to attend a clinic and may not have wanted to arrange for a courier. Six hundred and eighty-two (of 2,780) women did not have the full six months, although they were no more than 49 days short of six months.

In the randomised phase, 2/100 women in the clinic group were positive for HPV16 and 6/100 were positive for ‘Other’ hrHPV types (Table 4). In the home-based group, 16/205 were positive for hrHPV ‘Other’, with two of those women also being positive for HPV18; in total, 24/305 (7.9%) of the women who self-sampled were hrHPV positive. At colposcopy follow-up, three women had high-grade histology results. One woman who was HPV18 & ‘Other’ positive had a histology result of high-grade squamous intraepithelial lesion/adenocarcinoma in situ (HSIL/AIS) and two women who were positive for hrHPV ‘Other’ had results of HSIL.

**Table 4:** Positive high-risk human papillomavirus results and follow-up results.

| Randomised group | Screening history | Age Range | HPV self-sample result (type(s) detected) | Cytology follow-up result | Colposcopy follow-up result |
| --- | --- | --- | --- | --- | --- |
| Clinic | Never | 30-39 | 16 | Unsatisfactory | LSIL |
| Clinic | Under | 40-49 | 16 | ASC-US | LSIL |
| Home | Under | 30-39 | 18 & Other | Atypical Cells/HSIL/CIN2/3 | HSIL/AIS |
| Home | Under | 30-39 | 18 & Other | Negative | LSIL |
| Home | Under | 50-59 | Other | Negative | N/A |
| Home | Never | 40-49 | Other | Negative | N/A |
| Home | Never | 30-39 | Other | Negative | N/A |
| Clinic | Never | 40-49 | Other | Negative | N/A |
| Home | Never | 30-39 | Other | Negative | N/A |
| Home | Under | 30-39 | Other | Negative | N/A |
| Home | Never | 30-39 | Other | Negative | N/A |
| Home | Never | 40-49 | Other | Negative | N/A |
| Clinic | Never | 40-49 | Other | Negative | N/A |
| Home | Never | 40-49 | Other | Negative | N/A |
| Home | Never | 50-59 | Other | Negative | N/A |
| Home | Under | 60-69 | Other | ASC-US | N/A |
| Home | Never | 30-39 | Other | ASC-US | N/A |
| Home | Under | 30-39 | Other | LSIL/CIN1/HPV | Negative |
| Home | Never | 30-39 | Other | LSIL/CIN1/HPV | Negative |
| Clinic | Under- | 30-39 | Other | LSIL/CIN1/HPV | Immature squamous metaplasia |
| Clinic | Under | 40-49 | Other | HSIL suspicious for invasion | HSIL |
| Home | Never | 30-39 | Other | Atypical Cells/HSIL/CIN2/3 | HSIL/CIN3 |
| Clinic | Never | 50-59 | Other | Not completed | Declined follow-up |
| Clinic | Under | 40-49 | Other | Not completed | Lost to follow-up |

HPV: human papillomavirus; never: never screened; LSIL: low-grade squamous intraepithelial lesion; Under: under-screened; ASC-US: atypical squamous cells of undetermined significance; HSIL: high-grade squamous intraepithelial lesion; CIN2/3: cervical intraepithelial neoplasia 2 or 3; AIS: adenocarcinoma in situ; N/A: not applicable; CIN1: cervical intraepithelial neoplasia 1.
Two women in the home-based group had an invalid/equivocal result and did not provide a second sample. Another woman in the home-based group had an invalid/equivocal result and provided a second sample (which was negative for high-risk human papillomavirus). A fourth woman randomised to the home-based group had an invalid/equivocal result but took her sample in the clinic during the opportunistic phase. She did not provide a second sample.

Five of the 14 women in the usual-care group had normal cytology. Nine of the women randomised to usual care actually took an HPV self-sample rather than having a cytology sample taken by their healthcare professional; all of these samples were negative for hrHPV. None of the cytology results in the usual-care group was ‘unsatisfactory’ and none of the hrHPV results in the usual-care group was invalid/equivocal.

Follow-up was completed for 22/24 women (92%) who tested positive for hrHPV. One woman made an informed decision to decline follow-up after a shared decision-making conversation and the other was not contactable despite repeated attempts. The mean nurse-time required to achieve follow-up was 2.5 hours.

The laboratory turnaround time for the HPV samples received during the main phase of the study was a mean of 1.23 days, with a range of 0–18 days. Only 1/314 samples exceeded the expected 7-day turnaround (18 days) due to the Christmas/New Year shutdown period.

## Discussion

Sending women a self-sample kit for hrHPV testing at home resulted in statistically significantly (p≤0.001) higher participation than an invitation to have a usual-care cytology sample in the clinic among Māori (14.6% *vs*. 2.0%) and Asian (18.5% *vs*. 4.5%) and, to a lesser extent among Pacific women (8.8% *vs*. 1.7%). In the fully adjusted models, Māori women randomised to the home-based group were almost ten times more likely to participate than usual-care women (OR 9.7, 95%CI 3.0-31.5); Pacific women were six times more likely (6.0, 1.8-19.5) and Asian women around five times more likely (5.1, 2.4-10.9). This study is the first in Aotearoa New Zealand that specifically tested a mail-out approach for self-sampling for cervical-cancer screening and an intensive nurse-support model to achieve high rates of follow-up.

An invitation to take a self-sample at a clinic was less effective, suggesting that at least some of the barriers to clinic attendance are the same whether for self-sampling or a healthcare-professional-taken sample. Known opportunity costs and barriers to clinic attendance for cervical screening may explain why a higher proportion of home-based women participated^18^; they may also explain why the home-based group participated earlier than the clinic-based and usual-care groups.

The participation in our study was lower than that in a recent meta-analysis, which showed a pooled average intention-to-treat participation in the self-sampler mail-out group of 24.8%, compared to 11.5% in those who received a routine invitation or reminder strategies^2^. In contrast, the relative increase in participation was higher in our study *i*.*e*., uptake in the home-based group was almost five times that of the usual-care group (when comparing crude percentages of uptake between groups). Compared to the iPap trial^19^ in Australia, which involved women from the general population, did not include a clinic-based self-sampling group, and had a 6-month time period during which women could participate (compared to our 3-month period), our study had lower participation: 15% of never-screened and 14% of under-screened women, compared to 20% and 12%, respectively, in the iPap trial. Our study also had a lower participation in the usual-care group: 3% among never- and under-screened, compared to 6% and 6% in the iPap trial^19^. One likely contributor to the lower absolute participation observed in our study may have been our target population, which was intentionally focused on those women least served in the current health system: nondominant population groups and women living in areas of high socioeconomic deprivation, known to experience substantial barriers to care and differences in quality across the screening, diagnostic, and treatment pathway^11,20,21^. In addition, the higher participation in the iPap trial may be due to the longer time period during which the women could participate, although women in our study were sent a reminder letter/text, which did not occur in the iPap trial^22^.

A recent community-based cluster-randomised controlled trial (*He Tapu Te Whare Tangata*) in Aotearoa New Zealand, which involved intense recruitment of all overdue women, found that 59.0% of Māori women took up the face-to-face offer to self-sample compared to 21.8% who attended for a smear, with an adjusted risk ratio of 2.8 (2.4-3.1)^13^. The absolute participation proportions were therefore considerably higher; however, the odds ratio (reflecting a relative difference) was lower than that reported in our study (OR 9.7, 95%CI 3.0-31.5). The differences in participation may be due to different recruitment approaches: MacDonald *et al*^13^ used texting, email, letters, phone calls, and outreach services (nurses and kaiāwhina (non-clinical community Māori health workers)) to invite women to screening. As with the iPAP study, there are also likely to be differences in the target populations – overdue *vs*. our never-screened and highly under-screened populations and no European participants in our study. Population approaches, such as ours, are known to achieve lower participation than individual approaches^2^ but they do mimic the recruitment methods of routine screening. The degree of non-contactability (women not receiving an invitation) is unknown in our study. Our unpublished feasibility work revealed a very high proportion of non-contactable women (up to 50%) among a similar group, suggesting that participation may be substantially underestimated. As we expect that the proportion of non-contactable women is the same for the different arms of the trial, this should not affect the relative differences across groups.

In our study, 7.6% of the women who took a self-sample were positive for a hrHPV type, similar to the proportion in the iPap trial (8.5%)^19^, but lower than that in the *He Tapu Te Whare Tangata* trial (11.0%)^13^ (the latter trial included younger women known to have higher HPV prevalence). Our positivity proportion is at the lower end of the range found in the above-mentioned recent meta-analysis (6.0–29.4%)^2^. Despite the modest overall participation, the identification of clinically significant disease (Table 4) amongst HPV positive women from underserved groups is important to address known inequities in cervical cancer incidence and mortality.

During the six-month follow-on non-randomised sub-study, 192 (of 2,780; 6.9%) additional women self-sampled, suggesting that offering women home kits and self-sampling when they present for other reasons may also modestly increase screening uptake. It is notable that the women randomised to usual care took up the subsequent self-sampling opportunity more enthusiastically than those originally offered self-sampling. As with other forms of opportunistic invitation, it is important to ensure that women do not feel unduly pressured to be screened.

Clinically appropriate follow-up was discussed with all women with a positive HPV test (n=24) and was achieved in 92% (n=22). Both women who were not followed up have a recall notice in the practice management system for discussion and the offer of related future follow-up. Follow-up was higher in our study than in the trial by MacDonald *et al*^13^ (78%) and the iPAP trial^19^ (62%). Most HPV-positive women (18/24; 75%) needed a short interval of phone support to achieve clinically appropriate follow-up; however, 5/24 women (20%) needed additional support (*e*.*g*., a home visit for a follow-up cytology test) and 4% (1/24) of women needed very intensive support (*e*.*g*., multiple phone-calls and visits to discuss options plus transport and support to attend colposcopy).

A strength of our study was that we robustly tested different primary-care invitation approaches, which are potential policy options in Aotearoa New Zealand; this enabled us to assess the effects of different elements of the invitation approaches. Previous work had suggested that Māori women would respond better to face-to-face offers,^23^ although recent open community discussions indicated that Māori women would take up the offer of a mailed kit^18^.

The limitations of our study include the inability to ascertain whether we had the correct address or phone number for all women who appeared to be non-responders, a known limitation of population-recruitment approaches. Participation may therefore be underestimated. We invited women only in the Auckland and Waitematā DHB areas, so our findings may not be generalisable to women outside of those areas (*e*.*g*., rural areas). We were unable to blind participants and researchers to study-group allocation. Participants were aware that the study had three arms but not all of the details of home *vs*. clinic as the sites at which self-sampling would occur.

Based on feedback from the participating clinics, it may be that our participation is an underestimate of potential uptake if self-sampling were to be integrated into the NCSP because, in the setting of the study, there was a need to provide a large amount of trial-specific information; reading all this may have been a barrier, particularly for women for whom English is a second language and those with low health literacy. Our results thus reveal the lower bound of participation if self-sampling were introduced in Aotearoa New Zealand.

## Conclusion

The results of our study show that offering the opportunity to self-sample to underserved women who are also Māori, Pacific, and Asian, in Aotearoa New Zealand is likely to increase participation in cervical-cancer screening, particularly when mailed-out and completed at home.

## Supporting information

CONSORT Checklist Potter et al

Author COI Portfolio Potter et al

## Data Availability

As a result of ethics requirements and issues around data sovereignty associated with Indigenous people, at present we are unable to share data.

## Acknowledgements

We thank the staff of the participating Primary Health Organisations and GP clinics, and all of the women who took part in our study. We thank Marion Saville and Susan Reid for assistance with the design of the study materials, and Mellissa Murray for assistance with project administration and coordination. Thanks to Deralie Flower, lead colposcopist at Auckland DHB, and Amy Tam, Krish Pillay and the other staff at Anatomic Pathology Service. We also thank Prachee Gokhale at the Research Centre for Hauora and Health, Massey University for laboratory assistance. Thanks are also due to the cultural advisors Samantha Bennett, Leani Sandford, and Sulu Samu, and Waitematā DHB Asian Health Services who were available to take queries from participating women.

---

**Supplementary Table 1:** Exclusions by reason and ethnic group.

| Ethnic Group | Reason for exclusion |  |  |  |  |  |  |  |  |  |  |  | Uncontactable (after exclusions) | Opted out (after exclusions & uncontactable) | Incorrect address (after exclusions & uncontactable & opted out) | Screened outside of study (after exclusions & uncontactable & opted out) |
| --- | --- | --- | --- | --- | --- | --- | --- | --- | --- | --- | --- | --- | --- | --- | --- | --- |
|  | Deceased | History of high-grade smear result and not completed test of cure | Not active in clinic | Other | Previous history of or currently has cervical cancer | Has had a recent smear | Hysterectomy | Not eligible for NZ health services | Pregnant | Symptomatic | Participated in feasibility study | Total |  |  |  |  |
| Māori | 6 | 63 | 118 | 57 | 2 | 29 | 241 | 0 | 23 | 7 | 5 | 551 | 10 | 25 | 32 | 1 |
| Pacific | 2 | 27 | 176 | 63 | 1 | 63 | 162 | 5 | 20 | 4 | 0 | 523 | 6 | 29 | 35 | 6 |
| Asian | 0 | 15 | 252 | 33 | 0 | 65 | 230 | 29 | 39 | 3 | 0 | 666 | 9 | 58 | 36 | 6 |
| **Total** | 8 | 105 | 546 | 153 | 3 | 157 | 633 | 34 | 82 | 14 | 5 | 1,740 | 25 | 112 | 103 | 13 |

